# Assisted reproductive technology (ART) patient information-seeking behavior: a qualitative study

**DOI:** 10.1101/2023.12.07.23299684

**Authors:** Emma Mayette, Ariel Scalise, Angela Li, Nicolette McGeorge, Kaitlyn James, Shruthi Mahalingaiah

## Abstract

**Objective:** To investigate patient interaction with information sources while undergoing assisted reproductive treatment.

**Methods:** Semi-structured interviews with fifteen individuals were conducted between August and October 2022. Interview participants underwent assisted reproductive treatment including embryo transfer between January 2017 and April 2022 within a large urban healthcare system. Thematic analysis of the interview transcripts was performed.

**Results:** Participants reported that they engaged in informed decision-making with their provider. Three main themes were identified. Participants 1) utilized clinic-provided information and then turned to outside sources to fill knowledge gaps; 2) struggled to learn about costs and insurance; 3) had difficulty identifying mental health resources to support care.

**Conclusion:** Patients prefer clinic-provided resources and then utilize academic sources, the internet, and social media when they have unfulfilled information needs. Knowledge gaps related to cost, insurance, and mental health support were identified.

**Practice Implications:** ART clinics should consider providing more information about cost, insurance, and mental health support to patients.

## 1. Introduction

Approximately 13% of women in the United States of reproductive age seek infertility services [1]. Treatment options include ovulation-stimulating medication, artificial insemination, or assisted reproductive technology (ART) which includes in-vitro fertilization (IVF) [2]. IVF involves fertilization of extracted eggs with sperm in the laboratory and transfer of the fertilized embryo to the uterus [3]. This treatment can result in pregnancy and a live birth if successful.

Many people are unfamiliar with fertility healthcare before becoming patients [4]. Patients often possess large knowledge gaps about treatment and may have difficulty understanding ART care [5]. Patients receiving infertility treatment have reported seeking additional information on the internet to supplement their knowledge [6], [7]. In one study, 87.8% of respondents turned to the internet for additional infertility information, and 29.1% reported that they still did not find the answers they were looking for after searching online [8]. It is imperative to supply infertility patients with sufficient information resources so that they can make informed decisions about care [9].

Qualitative research can answer questions of clinical relevance by providing a thorough account of patient experiences [10]. In this study, semi-structured interviews were used to understand patient perceptions of the health information they were provided during ART care, as well as ways in which they interacted with different resources. The goal of this analysis was to investigate patients’ perceived ability to utilize information resources to understand ART treatment while undergoing care.

There are few existing studies in the United States regarding patient information-seeking behavior during ART care. Investigating the underlying complexities behind healthcare information resource utilization is an important first step towards supporting patient understanding of their ART treatment.

## 2 Materials and Methods

### 2.1 Ethical Approval

The Massachusetts General Hospital Institutional Review Board approved this study (#2022P000474) and informed consent was obtained from each participant.

### 2.2 Participants

Participants were identified from patient records at a large urban healthcare system. Eligible patients were those who underwent ART, including embryo transfer, between July 2017 and April 2022. The study participants were selected through purposeful sampling which allowed for the assessment of information-seeking behavior of patients that underwent ART care [11]. Semi-structured interviews were used to conduct an in-depth exploration of each participant’s knowledge and experience [12]. Patients interested in being contacted for research were invited to participate in the Assisted Reproductive Technology Patient and Provider Resource to Improve Communication about Outcomes and Treatments (APRICOT) study which included both a survey and an interview. Participants who completed the survey and indicated interest in an interview were contacted to participate. Participants were selected for the interview in the order that they completed the survey, with focus placed on recruiting a sample representative of the racial and ethnic distribution of the population of Massachusetts [13]. Interview participants were provided with $50 Target gift cards as compensation for completing the interview.

### 2.3 Data Collection

A semi-structured interview guide was developed using a human factors engineering-based approach which aims to assess and optimize the interaction between humans and elements of their environment [14]. This approach was selected to uncover the experiences of fertility patients with all aspects of their care including communication, information accessibility, and decision-making processes. The interview guide contained open-ended questions and participants were asked probing questions about topics they introduced [15]. The semi-structured guide was updated throughout the interview period to enhance clarity and ensure consistent probing. The complete interview guide is included in Supplementary Document 1. Video interviews via the healthcare system’s Health Insurance Portability and Accountability Act (HIPAA)-compliant Microsoft Teams (n=12) and telephone interviews (n=3) were conducted by an undergraduate research assistant (E.M.) with support from a PhD-level scientist trained in qualitative methods and human factors engineering (N.M.) between August 2022 and October 2022. E.M. was trained in qualitative techniques by A.S., who holds an MPH and has experience conducting qualitative interviews. The interview audio was recorded using a digital voice recorder for all interviews and transcribed verbatim via a HIPAA-compliant professional transcription service. The audio files were deleted after transcription was completed. The interviews ranged from 41 to 60 minutes in length with a mean length of 50 minutes. The final sample size was 15 participants because this was the point at which data saturation was reached and no new ideas were brought up by participants in response to the interview guide questions.

### 2.4 Data Analysis

Prior to conducting analysis, all personal identifying information was removed from the transcripts. An inductive approach was used to identify interview themes, meaning the analysis was done without applying an a priori hypothesis [16]. Thematic analysis was performed according to the six phases developed by Braun and Clarke: familiarizing yourself with your data, generating initial codes, searching for themes, reviewing themes, defining and naming themes, and producing the report [17]. Researchers familiarized themselves with the data by reading the interview transcripts. Initial code generation was performed by E.M. and A.L. who each individually analyzed one interview using NVivo 12.0 software [18]. A.L. is a resident physician in obstetrics and gynecology and was an MPH student at the time of this study. After reviewing an interview transcript, researchers met to compare and combine their coding until they reached agreement for five interviews. A third reviewer (A.S.) contributed to the decision-making process when disagreement occurred. A codebook containing the list of codes and definitions was created (Supplementary Document 2). The coders then each individually analyzed the remaining ten interviews, with E.M and A.L. each completing five interviews. Once coding was complete, E.M. and A.L. reviewed the coding performed by the other researcher to ensure consistency. To begin searching for themes, codes related to patient information-seeking behavior were sorted into preliminary groups to organize the data. Preliminary themes containing robust support across the fifteen interviews were refined to ensure they contained concise ideas. The data extracts associated with each theme were reviewed to ensure they formed meaningful patterns that accurately represented the dataset. Researchers then reviewed compiled themes before defining and naming final themes. The final report writing was informed by the recommendations included in the Standards for Reporting Qualitative Research (SRQR) [19].

## 3. Results

The median age of participants at the time of interview was 39 years. Most participants had a live birth after treatment (73.3%). Nearly all participants completed college or held post-graduate degrees (93.3%). The demographic information of interview participants is detailed in Table 1.

**TABLE 1:** Characteristics of Participants (N = 15)

| Characteristic | Mean (SD) |
| --- | --- |
| *Age in years* | 39 (0.50) |
|  | N (%) |
| *Biological sex* |  |
| Female | 15 (100%) |
| *Race/ethnicity* |  |
| White | 8 (53.3%) |
| Hispanic, Latina, or Spanish Origin | 2 (13.3%) |
| Black, African American, or African | 1 (6.7%) |
| Southeast Asian | 1 (6.7%) |
| South Asian | 1 (6.7%) |
| Some other race, ethnicity, or origin | 1 (6.7%) |
| *Education* |  |
| Some college | 1 (6.7%) |
| College graduate | 5 (33.3%) |
| Postgraduate (for example masters, professional, doctorate) | 9 (60%) |
| *Outcome of most recent ART cycle* |  |
| Live birth | 11 (73.3%) |
| Spontaneous abortion | 1 (6.7%) |
| Outcome unknown | 3 (20.0%) |

Participants in the semi-structured interviews discussed feeling content with their medical care when they fully understood their treatment. Participants utilized outside information sources to learn about their ART care and further engage with their providers: *“I mostly just let the clinic tell me what the protocol was going to be, and then I kind of read a little bit more about it to make sure that it made sense for me. I think I ultimately just agreed with what they decided anyways, but at least it made me feel like … I could have at least gone back and asked why they chose that or why not something else and had the conversation*.*” – Participant 9*. The participants felt confident in the treatment plans created by their providers after they supplemented their knowledge with additional resources.

Three main themes were identified (Figure 1). Interview participants: 1) utilized clinic-provided information and then turned to outside sources to fill knowledge gaps; 2) struggled to learn about costs and insurance; 3) had difficulty identifying mental health resources to support care.

**Figure 1.**
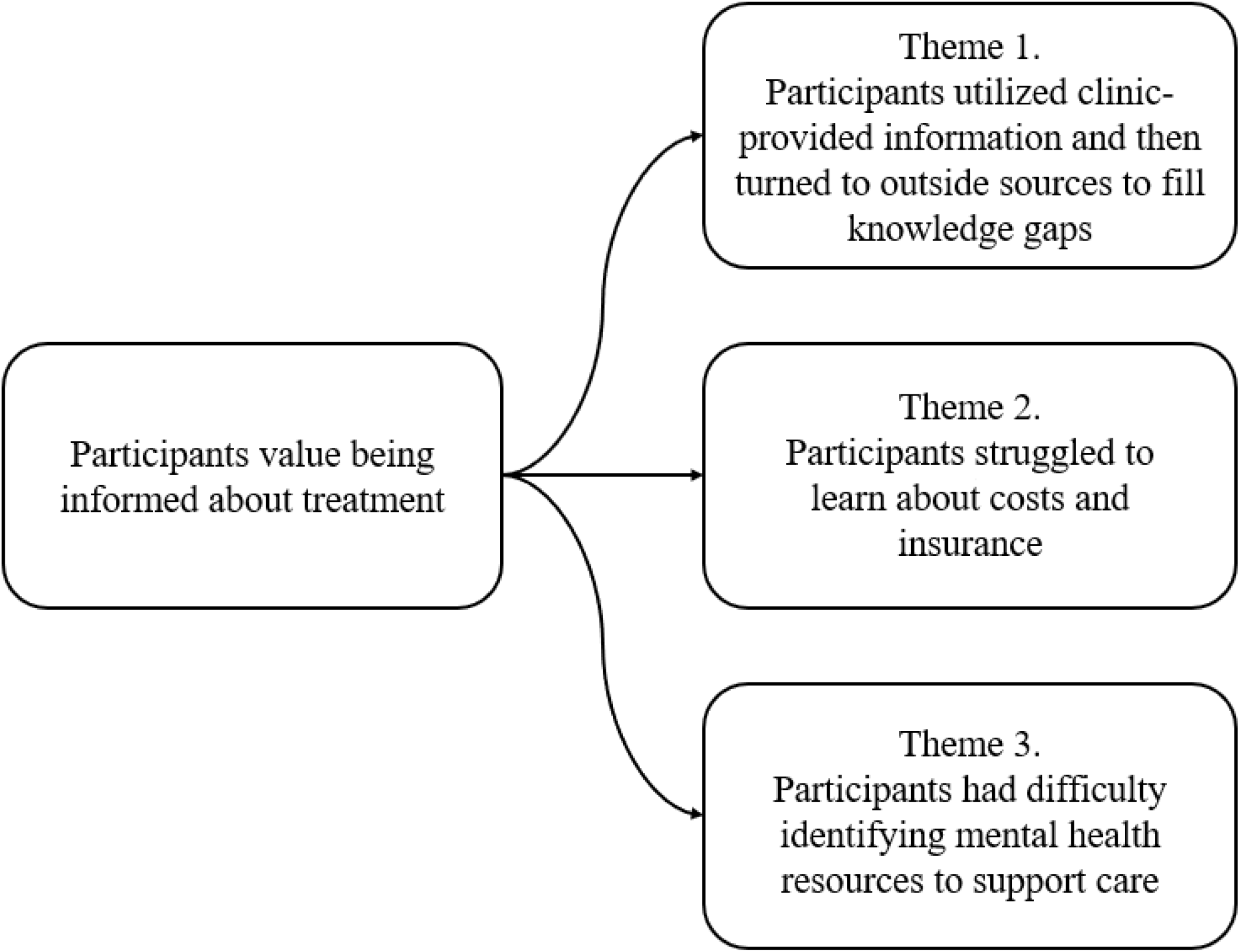
Summary of results.

### Theme 1 Participants utilized clinic-provided information and then turned to outside sources to fill knowledge gaps

Participants reported using four categories of resources: clinic materials, academic sources, general internet searches, and social media. Participants reported a preference for information from their clinic and provider. They supplemented their knowledge with outside sources such as academic websites. They lastly reported searching the internet and social media for further information that they could more easily understand.

#### A. Clinic materials

Participants spoke positively about the quality of information they received from their clinic and that they engaged with these sources prior to searching on their own. Participants reported a preference for receiving information from their providers:

> *“I don’t like to search on my own, I’d rather just talk to a doctor*.*” – Participant 12*

Participants reported using clinic-provided materials; most notably they watched videos about how to administer medication:

> *“I loved how they had the videos on how to do the medication, and those were up whenever you could watch them*.*” – Participant 1*

#### B. Academic sources

When participants wanted to find more information, they reported turning to sources such as academic organizations and research studies. However, they reported difficulty understanding the information provided by these sources.

> *“I was checking everywhere and anywhere. College resource pages…had some papers on stuff. But I don’t feel like it made me any smarter about what I was doing*.***”*** *– Participant 10*

#### C. Internet searches

Next, participants reported performing general internet searches to find answers:

> *“I did google things, probably a little more than I should have, and I think that can lead you down paths that are not helpful”-Participant 13*

Participants reported awareness that internet resources can lead to misinformation and heighten stress:

> *“I actually ended up researching on the internet and all of that, there were a lot of negative things around how people’s experiences were. That scared me a lot*… *I thought, ‘Okay. I’m not going back--I’m not going to research*.*’” – Participant 14*

#### D. Social media

Participants utilized social media sites to connect with other fertility patients and find more information about care:

> *“So after I saw the doctor, I joined a Facebook group, and I think I learned a lot, actually, through that, just from reading other people’s questions and hearing other people’s stories, and posting my own questions on there. So I think most of the information I actually got was through that” – Participant 7*

Participants utilized these sources to learn more about treatment, particularly anecdotal information:

> *“For example, on the patient education video, they told you to put one thing in your thigh, but when I did that, I would keep hitting veins, and I had huge bruises. So then I looked online, and they all recommended a different spot in your stomach. So there were just suggestions in terms of the gauge of the needle, but that wasn’t covered in the education stuff. That was really helpful. And putting a cold compress on to numb it. That sort of thing*.*” – Participant 3*

### Theme 2 Participants struggled to learn about ART costs and insurance

Participants reported that they did not know how insurance coverage would apply to their care, as well as how much this type of care might cost. Furthermore, patients reported trouble finding information about ART costs and insurance:

> *“Oh, my gosh. Just figuring out how many cycles. So my husband’s insurance only covered, I think, it was two IVF cycles and $10,000 worth of medication. I didn’t know that, basically, one cycle is like $11,000 or $11,500 in medications, so that freaked me out*.*” – Participant 1*

### Theme 3 Participants had difficulty identifying mental health resources to support ART care

Many interview participants reported high levels of emotional strain while they were undergoing ART treatment:

> *“I think the whole process is just emotionally gut-wrenching. You’re kind of a basket case the whole time*.*” – Participant 2*

Some participants further reported lacking the tools necessary to find mental healthcare. Participants reported that they did not know about mental healthcare options at the time and wished they had sought mental health support due to the strenuous nature of treatment:

> *“I probably should’ve seen some kind of mental health--maybe just a counselor or a therapist, just to check in and see how I was doing, just because those were really, really dark times for me… Unfortunately, it wasn’t something easy to find*.*” - Participant 11*

## 4. Discussion and Conclusion

### 4.1 Discussion

This study assessed the information-seeking experiences of fertility patients through a qualitative, patient-centered lens. Participants indicated that they value informed decision-making, prefer clinic-provided materials, and desire more information regarding cost, insurance, and mental healthcare. As noted in other literature, healthcare patients value being informed about their care so that they feel confident about their treatment plan [20]. Interview participants reported the importance of informed decision-making, reiterating the significance of this analysis to identify the types of information ART patients currently lack.

In this study, participants reported a preference for information provided by their provider and clinic. They supplemented this knowledge with outside sources, including academic websites, research studies, internet searches, and social media. These findings are consistent with a previous study in which patients reported searching the internet for information about care but valued their physician’s opinion over outside sources [21].

Utilizing social media for health information has been observed in patients with unfulfilled information needs [22]. OB-GYN patients have reported using social media for education, social support, and sharing of advice, but were aware that these sources can contain inaccurate information [23]. Social media can be a beneficial tool for healthcare patients to share advice and receive community support [24]. However, while social media enables connection with others undergoing similar experiences with fertility care, it is not a trusted tool for reliable information about treatment [25]. There is a large amount of misinformation on general internet websites, including social media websites, which can be detrimental to accurate patient knowledge about health [26]. Providing patients with validated and easily accessible information can help prevent misinformation from unregulated sources, as well as decrease the amount of time providers spend educating patients.

The desire for more information about insurance and cost of treatment is true for patients across healthcare disciplines in the United States [27]. It is expected that individuals will have difficulty locating and navigating insurance policy information [28]. Fertility coverage is difficult to navigate because of varying insurance laws and regulations in different states [29]. The need for accurate insurance and cost information is imperative as insurance coverage for ART varies widely across and within states [30]. While insurance coverage is different for every patient, fertility clinics should provide guidelines on obtaining this information due to reported complexities in this area. Providing patients with cost estimates of fertility treatments prior to care can improve their ability to make informed decisions [31].

We found that interview participants experienced high levels of emotional distress during their fertility care and had difficulty identifying mental health support options. Fertility patient struggles with mental health and the limited provision of information regarding psychological support tools has been previously noted [32]. In one survey, fertility patients responded that they were not adequately informed about social support options during fertility care [33]. Meeting the psychosocial needs of fertility patients is an important aspect of patient-centered care [34]. Given the high rates of depression and anxiety experienced by patients undergoing ART [35], providers should ensure that patients are aware of mental health resources available to them, as well as the potential benefits of their utilization.

This study has several limitations. Most participants had a live birth, which may have influenced willingness to participate in the study and the perception of their care experience. Nearly all participants had completed college or professional degrees, which is not representative of the United States population. The participants received care in a state that mandates insurance coverage for ART care, but there are many states throughout the U.S. that do not ensure coverage [36]. The study sample includes patients from a single health care system that has extensive clinical resources. Therefore, the knowledge gaps reported in this study may potentially be further exacerbated at clinics with fewer resources.

ART technology is rapidly evolving and education resources must be updated to keep patients informed about their fertility treatment [37]. ART treatment, laws, and regulations can vary greatly between countries based on cultural and economic factors [38]. The results from this study can be used to inform clinics in the United States about ways in which patients interact with information sources to learn more about ART treatment and existing patient knowledge gaps.

### 4.2 Conclusion

This study qualitatively assessed the information-seeking experience of patients undergoing ART treatment. Patients reported a preference for utilizing clinic-provided information sources to learn more about care. They next turned to academic sources, which they had difficulty understanding. Lastly, they utilized the internet and social media sites to gain more information. Patients had difficulty learning about ART costs, insurance, and support for mental health during treatment.

### 4.3 Practice Implications

ART clinics should consider providing patients with more information related to cost, insurance, and mental health support tools tailored to a non-clinical audience to ensure understanding.

## Data Availability

De-identified data can possibly be made available upon reasonable request.

## Author Contributions

**Emma Mayette:** Writing – original draft, Formal analysis, Investigation, Data curation, Writing – review & editing, Project administration, Methodology. **Ariel Scalise:** Conceptualization, Methodology, Formal analysis, Investigation, Writing – review & editing, Project administration, Funding acquisition. **Angela Li:** Formal analysis, Writing – editing & reviewing. **Nicolette McGeorge:** Conceptualization, Methodology, Investigation, Writing – review & editing, Funding acquisition. **Kaitlyn James:** Formal analysis, Methodology, Writing – review & editing. **Shruthi Mahalingaiah:** Conceptualization, Methodology, Writing – review & editing, Supervision, Funding acquisition

## Acknowledgements

We gratefully thank Shoba Ramanadhan and Rachel Vanderkruik for providing their qualitative expertise during the editing process.

## Funding

This project is supported by the Centers for Disease Control and Prevention of the U.S. Department of Health and Human Services (HHS). The contents are those of the authors and do not necessarily represent the official views of, nor an endorsement, by CDC/HHS, or the U.S. Government.

## Declaration of Competing Interest

The authors disclose no competing interests.

## Data Availability

De-identified data can possibly be made available upon reasonable request.

## Notes

### Competing Interest Statement

The authors have declared no competing interest.

### Author Declarations

IRB of Massachusetts General Hospital gave ethical approval for this work

