## Supplementary Document 1 for "Assisted reproductive technology (ART) patient information-seeking behavior: a qualitative study"

Supplementary Document 1. Semi-structured interview guide

- Do you mind briefly describing your IVF journey to me?
  - Where do you receive your care?
  - What was your main diagnosis (male factor, ovulatory, PCOS, endometriosis, tubal factor, fibroids)?
  - What treatments have you received?
  - Where are you in your treatment?
  - What was your outcome?
- How was your overall experience with fertility treatment?
  - What were/are some positive aspects of your journey?
  - What about your journey was/is challenging?
- Were there any specific challenges associated with your clinic? Ex. access to information, communication with doctor
- How did you begin your journey with your infertility doctor?
  - How did you learn about different IVF clinics?
  - What factors lead you to choose your current clinic?
- What resources do you use related to your infertility journey?
  - Websites or sources you searched on your own?
  - Protocols to follow or info from doctor?
  - How did you find these? How did you stay organized during your care?
  - What type of information were you searching for? What type of information did you need when you first got started? During or after treatment?
- What tools or technologies do you use related to your infertility journey?
  - Phone apps, social media (Instagram, YouTube, etc.)
    - Apps: how does it function, what do you use it for, how did you find it
  - What about those tools works well?
  - What about using those tools is challenging?
  - Patient gateway app clinic
- What about technology use for other types of healthcare?
- How did you communicate with your clinic during ART treatment? After treatment?
  - Did you share information with your clinic after you graduated from ART care?
  - Who initiated this contact? (Patient or clinic)
  - Which member of your team did you communicate with regarding your outcomes?
  - How did your clinic contact you?
  - What about this process worked well? What was challenging?
  - What type of information did you share with your clinic?
- How did you communicate with family and friends about your ART treatment?
- Would you recommend IVF treatment to family or friends if they were struggling with fertility?
- Have you ever looked at public websites that report national IVF data such as pregnancy rate, multiples rate, or used a tool to calculate your success rate?
  - Which one?
  - How did you know to look there?
  - What did you learn?
  - Have you heard of ART CDC mandated reporting?
  - Do you know what the purpose of mandated reporting of ART outcomes is?
- The purpose of this survey is to learn about patient experiences reporting their outcomes back to make patient experiences smoother. How do you think a phone app could improve IVF outcomes reporting experiences for you as a patient?
- Do you have any thoughts about what app features might be useful to you?
- Lastly, I would like to ask if there is anything we didn’t get a chance to talk about yet that you wish to share regarding your IVF treatment?
