## Supplementary Document 2 for "Assisted reproductive technology (ART) patient information-seeking behavior: a qualitative study"

Supplementary Document 2. Qualitative codebook

| **Code** | **Description** |
| --- | --- |
| Access to Care | Avenues through which patients first accessed or were introduced to fertility care |
| Clinic Reputation | Patient accessed care at their fertility clinic due to clinic reputation |
| Provider Recommendation | Patient accessed care at their fertility clinic due to recommendation by a healthcare provider |
| Difficult for Care | Factors that were unhelpful during the patient’s fertility care and/or made their experience difficult NOTE: Include all general references to other miscellaneous things that patients reported were difficult/unhelpful during their care that do not fit into an existing child code |
| Care Partner Issues | Difficulty in care due to: Issues related to the patient’s care partner (ie. spouse, parent, friend, etc.) |
| Commute to Clinic | Difficulty in care due to: Distance or time required for patient and/or partner commute from home to clinic |
| Complexity of Investigations & Treatment | Difficulty in care due to: Amount, frequency, or complexity of fertility-related investigations (bloodwork, ultrasounds, etc) and treatments (medications, procedures, etc) |
| Cost & Insurance Coverage | Difficulty in care due to: Issues related to the cost of fertility treatment or obtaining insurance coverage |
| COVID | Difficulty in care due to: The impacts of COVID and/or COVID restrictions |
| Delayed Referral to Care | Difficulty in care due to: Perceived or real delays in patient being referred to fertility care |
| Injections | Difficulty in care due to: Administration of injection medications during fertility treatment. Reasons may include pain/discomfort or coordination of injections. |
| Inter-Provider Miscommunication | Difficulty in care due to: Inaccurate or insufficient communication between healthcare providers |
| Lack of Communication | Difficulty in care due to: Inaccurate or insufficient communication between care provider and patient |
| Medication Issues | Difficulty in care due to: Issues related to medication administration, organization, supply, etc. |
| Multiple Providers | Difficulty in care due to: Receiving care from multiple different healthcare providers |
| Physical Discomfort | Difficulty in care due to: Physical discomfort or pain experienced during fertility investigations or treatments |
| Poor Provider Attitude | Difficulty in care due to: Issues related to the attitude of one or multiple of their healthcare provider(s) |
| Scheduling Issues | Difficulty in care due to: Issues related to the scheduling of clinical care (ie. appointments, treatments) resulting in delays or suboptimal timing of care |
| Side Effects | Difficulty in care due to: Clinical side effects resulting from investigations or treatments completed during the course of fertility care |
| Treatment Unpredictability | Difficulty in care due to: Issues related to the unpredictable nature of fertility treatments or unanticipated issues encountered during the course of treatment |
| Emotions During Treatment | Emotions experienced by the patient during the course of their fertility treatment |
| Anger & Frustration | Feelings of being upset, annoyed, angry, or frustrated |
| Distrustful | Feelings of a lack of trust or reassurance |
| Excitement & Joy | Feelings of enthusiasm, happiness, excitement, etc. |
| Nervous & Anxious | Feelings of worry, anxiety, unease, or nervousness |
| Non-Specific Emotional Strain | General non-specific comments pertaining to emotional strain, distress, or stress related to the fertility treatment process |
| Resolute Attitude | “Whatever it takes” attitude. Attitudes of being purposeful, determined, and unwavering during the process. |
| Sad & Depressed | Feelings of sorrow, grief, devastation, sadness, or depression |
| Helpful for Care | Factors that were helpful during the patient’s fertility care and/or improved their experience NOTE: Include all general references to other miscellaneous things that patients reported were helpful/beneficial during their care that do not fit into an existing child code |
| 1-to-1 Discussion | Improved experience due to: 1-to-1 individual discussions with care providers |
| Flexibility in Personal Life | Improved experience due to: Flexibility in patient’s personal life circumstances while undergoing fertility care |
| Geographical Location | Improved experience due to: Geographical location of the patient’s fertility clinic. Reasons include proximity to home, family/friends, other care facilities, etc. |
| Informed Decision Making | Improved experience due to: Adequate counselling and education from care providers/other sources to allow patient to make an informed decision regarding their care |
| Organization of Care | Improved experience due to: Aspects of care delivered in an efficient and organized manner |
| Positive Provider Attitude & Relationship | Improved experience due to: Positive attitudes and behaviors from care providers or reference to a good/positive relationship with their provider(s) |
| Referral to Other Specialists | Improved experience due to: Appropriate referral to other medical specialists as needed |
| Support Network | Improved experience due to: Psychological, social, and/or physical support from friends and family within patient’s network |
| Indication for Care | Reasons that patients were initially referred to fertility care |
| Anovulatory Condition | Reason that patient was referred to fertility care was: Infertility due to hormonal conditions that cause patient not to ovulate or to ovulate infrequently (ie. PCOS) |
| Genetic Testing | Reason that patient was referred to fertility care was: Primarily to undergo genetic testing of patient or embryos. Examples include extended carrier screening (ECS) or pre-implantation genetic testing (PGT) for monogenetic diseases (PGT-M) or aneuploidy (PGT-A). |
| Male Factor | Reason that patient was referred to fertility care was: Male factor causes of infertility (sperm/semen issues) |
| Maternal Age | Reason that patient was referred to fertility care was: Advanced maternal age (explicitly stated or implied) |
| Miscarriage | Reason that patient was referred to fertility care was: History of previous miscarriage (one or recurrent) |
| Prior to Surgery or Medical Treatment | Reason that patient was referred to fertility care was: To undergo fertility preservation prior to an anticipated medical (ie. chemotherapy) or surgical treatment for a separate medical issue |
| Tubal Factor | Reason that patient was referred to fertility care was: Tubal factor infertility (one or both blocked fallopian tubes) |
| Unexplained Infertility | Reason that patient was referred to fertility care was: Unknown or undiagnosed cause of infertility |
| Uterine Fibroids | Reason that patient was referred to fertility care was: Infertility related to uterine fibroids |
| Information Source | Resources from which patients are obtaining information related to fertility care NOTE: Patient must be referring to a resource they actually used themselves during fertility treatment (not hypothetical) |
| Academic & Professional Organizations | Information source: Academic and professional organizations (ie. medical association, hospitals/medical centers, universities, etc.) via. online website or printed material |
| Apps | Information source: Smartphone apps containing information related to fertility |
| Family & Friends | Information source: Provided from friends or family with previous experience in healthcare or fertility care |
| Fertility Clinic Materials | Information source: Informational and educational materials provided from the patient’s fertility clinic |
| Google | Information source: Google or other online search engine leading to utilization of misc. online websites (those not specified under another existing code). |
| Healthcare Admin | Information source: Administrators from fertility clinics, hospitals, or other medical centers |
| Healthcare Provider | Information source: Information provided directly from a healthcare provider (fertility doctor, other doctor, nurse, etc.) |
| News Articles | Information source: News articles that have been published in the general media |
| Online Forum | Information Source: Online forums tailored to people undergoing fertility treatment or prenatal care |
| Research Studies | Information Source: Published academic research studies |
| Social Media | Information Source: Social media posts |
| Patient Knowledge Gaps | Reported and perceived knowledge gaps by patients |
| Cost | Lack of knowledge around: Costs associated with fertility investigations and treatment |
| Embryo Transfer | Lack of knowledge around: Considerations and nuances related to embryo transfer (ie. number of embryos to transfer) |
| Infertility | Lack of knowledge around: Difficulty conceiving and areas related to fertility/infertility |
| Insurance | Lack of knowledge around: Insurance coverage of fertility treatment |
| Options After Failed Treatment | Lack of knowledge around: Options for patient to take following a failed treatment or IVF cycle |
| PGT | Lack of knowledge around: Pre-implantation genetic testing (PGT) for monogenetic diseases (PGT-M) or aneuploidy (PGT-A) |
| Risks & Complications of Treatment | Lack of knowledge around: Potential risks and complications due to fertility treatment (general or specific). Examples include: Ovarian hyper stimulation syndrome and other causes for IVF cycle cancellation, Multiple gestation (twins/triplets), etc. |
| Test Information | Lack of knowledge around: Information about fertility investigations including their risks, indications, accuracy, etc. |
| Treatment Information | Lack of knowledge around: Information about fertility treatments including risks, considerations, success rates, requirements, etc. |
| Patient Resource Ideas | All general questions and responses related to ideas and topics for a patient resource |
| Appointment Calendar | Calendar that keeps track of their clinic visits Example: “But it was a lot of keeping track of appointments and things like that. And so that would be another great thing to have in the portal. If you had a calendar that they could put your dates into with alerts and reminders and stuff like that, so they were able to put it in from their end, as well.” |
| Communicate with Providers | Ability to message with providers in the technology resource |
| Connect with Other Patients | Ability to connect with others undergoing IVF |
| Example Timelines | Schedules of ART care so that patients understand how long the process takes Example: “I feel like something with common overview of the process or next steps could be helpful as well. Because I know they told it to me. But when you're in it, I remember sometimes being like, "Okay. What happens after this? We do this for five days or how many days?" Just kind of seeing where it fits into the bigger picture could be helpful also to have.” |
| General ART information | Reliable information about ART care |
| How to Prepare for IVF | Things you should do prior to IVF so you don’t further extend your timeline |
| Information About Treatment Protocol | Example: “And then questions throughout the IVF process, that would have been amazing to have somewhere where you could look or even a portal that has a lot of resources that are specific to that treatment or that hospital or how they kind of do things, their process.” |
| Injection Tips | Tips on how to make injections easier |
| Medication Calendar | Calendar to keep track of days and times they need to take their medication |
| Mental Health Resources | Information about available mental health resources and how to get started with them Example: “But at the time, I don't remember being told, "Oh, here's someone you can talk to that can help you with this, this, and this, dealing with the mental health aspect of going through a process like this." I don't remember anyone ever offering or talking to me about that sort of thing, so I didn't seek it out at the time. But looking back, obviously, I wish that I had.” |
| Other Healthcare | References to aspects from other types of healthcare patient has accessed that they liked |
| Provider Profiles | Information/photos of other providers in the practice so they can learn about them before they might perform embryo transfers, etc. Example: “We wish we'd known those providers ahead of time. I guess that's part of it, too. You go in to have this transfer done or retrieval done, and it's kind of a big deal. And this person walks in you've never seen before to do everything, and you kind of wish you'd, at least, met that person beforehand.” |
| Success Rates | Success rates for clinic, nationally, so patients understand the probability of success |
| Test Results | Copies of test results easily accessible in the technology resource |
| Patient-Provider Communication | Methods used for communication between the patient and their care providers outside of in-person clinical visits NOTE: Must refer to a communication method actually used by the patient during their treatment, NOT hypothetical |
| Email Communication | Communication method used: Email |
| Patient Portal | Communication method used: Virtual patient portal (ie. Gateway) |
| Phone Communication | Communication method used: Phone calls |
| Pregnancy Outcome Reporting | All general questions and responses related to pregnancy outcome reporting |
| Treatment Organization | Methods used by patients to organize and schedule their fertility treatment |
| Alarms & Alerts | Organization method: Alarms and alerts to notify patient of medication administration times, appointments, etc. |
| Calendar | Communication method used: Calendar in any type of format (paper or virtual) |
| Fertility Apps | Communication method used: Smartphone apps designed specifically for fertility patients (ie. Tilly) |
| Paper Documents | Communication method used: Physical paper notes or documents |
| Virtual Documents | Communication method used: Virtual or online documents (ie. Google Doc, iPhone Notes app, Evernote, etc.) |
